# Baricitinib restrains the immune dysregulation in COVID-19 patients

**DOI:** 10.1101/2020.06.26.20135319

**Authors:** Vincenzo Bronte, Stefano Ugel, Elisa Tinazzi, Antonio Vella, Francesco De Sanctis, Stefania Canè, Veronica Batani, Rosalinda Trovato, Alessandra Fiore, Varvara Petrova, Francesca Hofer, Roza Maria Barouni, Chiara Musiu, Simone Caligola, Laura Pinton, Lorena Torroni, Enrico Polati, Katia Donadello, Simonetta Friso, Francesca Pizzolo, Manuela Iezzi, Federica Facciotti, Piergiuseppe Pelicci, Daniela Righetti, Paolo Bazzoni, Mariaelisa Rampudda, Andrea Comel, Walter Mosaner, Claudio Lunardi, Oliviero Olivieri

**Author notes:** Corresponding author. Prof. Vincenzo Bronte, Immunology Section, Department of Medicine, University and Hospital Trust of Verona. denotes equal contribution (Vincenzo Bronte, Stefano Ugel).

## Abstract

Severe acute respiratory syndrome coronavirus 2 (SARS-CoV-2) is the causative agent of the ongoing pandemic coronavirus disease 2019 (COVID-19). The majority of patients with COVID-19 have a good prognosis, but variable percentages in different countries develop pneumonia associated with lymphocytopenia and severe inflammatory response due to uncontrolled release of cytokines. These immune mediators are transcriptionally regulated by JAK-STAT molecular pathways, which can be disabled by small molecules. Here, we provide evidences on the efficacy of baricitinib, a JAK1/JAK2 inhibitor, in correcting the immune abnormalities observed in patients hospitalized with COVID-19. Indeed, we demonstrate a significant reduction in serum levels of interleukin (IL)-6, IL-1β and tumor necrosis factor (TNF)α, a rapid recovery in circulating T and B cell frequencies and an increased antibody production against SARS-CoV-2 spike protein in baricitinib-treated patients. Moreover, treated patients underwent a rapid reduction in oxygen flow need and progressive increase in the P/F. Our work provides the basis on developing effective treatments against COVID-19 pathogenesis using on-target therapy.

## Introduction

The pandemic spread of a novel, highly pathogenic coronavirus (SARS-CoV-2) has found the international medical community largely unprepared on prophylactic and therapeutic measures (1). The resulting syndrome, known as COVID-19, is characterized by a profound dysfunction of the upper and lower respiratory tract, with severity ranging from mild to moderate respiratory failure, up to acute respiratory distress syndrome (ARDS) that is generally fatal (2). Recently, the crucial role of the “cytokine release syndrome (CRS) (3), also referred to as “cytokine storm”, in acute lung damage and ARDS (4, 5) has become evident, thus providing the theoretical ground for therapeutic approaches able to interfere with the inflammatory cascade. Indeed, while the majority of patients either asymptomatic or with early stage of the disease are able to clear the infection, some patients with moderate disease, requiring hospital admittance, progress to a clinically severe phase associated with the “cytokine storm” within 10 days from symptom onset. These observations suggest that in some patients the immune response might be skewed and unable to neutralize the effects of the viral infection. For this reason, in addition to anti-viral therapy, immune modulators of cytokine production have been advanced.

Baricitinib is an oral, selective and reversible inhibitor of the Janus kinases JAK1 and JAK2, which was previously shown to dampen inflammatory immune responses and approved for indications such as rheumatoid arthritis (RA) (6). The drug was licensed at a daily dose of 2 mg/orally with good results in terms of clinical response and safety (7). In a recent meta-analysis, no statistically-significant increase in the risk of serious infections was recognized over a long treatment period (8), thus the use of this agent for a short 14 day period should have “trivial” adverse activity (9). In addition to the potential cytokine inhibitory activity, baricitinib was predicted to inhibit ACE-mediated endocytosis of SARS-CoV-2 by machine learning algorithms (9).

We hypothesized that JAK-STAT pathway inhibition might prevent the progression towards a severe/extreme form of the viral disease by modulating the patients’ immune response. Here we provide evidences that baricitinib-induced changes in the immune landscape were associated with a favorable clinical outcome of patients with COVID-19 pneumonia.

## Results and Discussion

### Baricitinib improves clinical parameters of SARS-CoV-2 infection

To understand the clinical impact of baricitinib to COVID-19, we assessed 20 patients who were admitted, within the period from March 18^th^ to April 18^th^ 2020, to the Unit of Internal Medicine at the University Hospital of Verona and Pederzoli Hospital of Peschiera with diagnosis of COVID-19 pneumonia, which was confirmed by the positivity of nasal swab to SARS-CoV-2 reverse-transcriptase-polymerase-chain-reaction assay.

In total, 88 patients (44 M/44 F) affected by COVID-19-related pneumonia were followed during the hospitalization. All subjects were treated with either hydroxicloroquine or antiviral therapy (lopinavir/ritonavir) as single agents or in combination (hydroxicloroquine plus antiviral therapy) according to clinical features. Supportive therapy, such as antibiotic prophylaxis and anticoagulant treatment, was provided at the discretion of the clinicians (Table S1). Steroids therapy was systematically avoided. Twelve (6 M/6 F) of these patients were excluded from the analysis considering their positive active history of malignancies: 2 hematological disorders (a multiple myeloma and an acute myeloid leukemia) and 10 cases of solid malignancies, including lung and breast cancers as well as kidney, prostate, ovarian and gastro-intestinal tumors. Arterial hypertension and cardiovascular disease as well as diabetes, chronic obstructive pulmonary disease, and chronic kidney disease were the prevalent morbidities in the other 76 subjects (Table S1). Among them, 20 subjects received the full course of baricitinib according to the study protocol. The other 56 subjects were considered as control group. According to the inclusion criteria and to baricitinib pharmacokinetics, patients were treated with baricitinib 4 mg twice daily for 2 days followed by 4 mg per day for the remaining 7 days. A low dose of 2 mg twice daily for 2 days followed by 2 mg per days was maintained for patients older than 75 years of age. A dose reduction was also considered in case of renal insufficiency (GFR < 30 ml/min/1.73 m^2^), hepatotoxicity or myelotoxicity.

Patients included in the baricitinib-treated group were matched with those included into the control group for age, sex, comorbidities and for several clinical features values (Table S1). Indeed, we did not observe differences in the symptoms ascribed to COVID-19, such as fever and cough, between the two patient cohorts. Moreover, patients in the two groups were clinically similar for several respiratory parameters, such as respiratory frequency, P/F ratio, and need of oxygen replacement therapy (Table 1), although baricitinib group showed a more severe radiologic score. Laboratory parameters were homogeneous among the two groups except for lactate dehydrogenase (higher in baricitinib-treated group) and D-dimer (lower in baricitinib-treated group, Table 1).

**Table 1.**
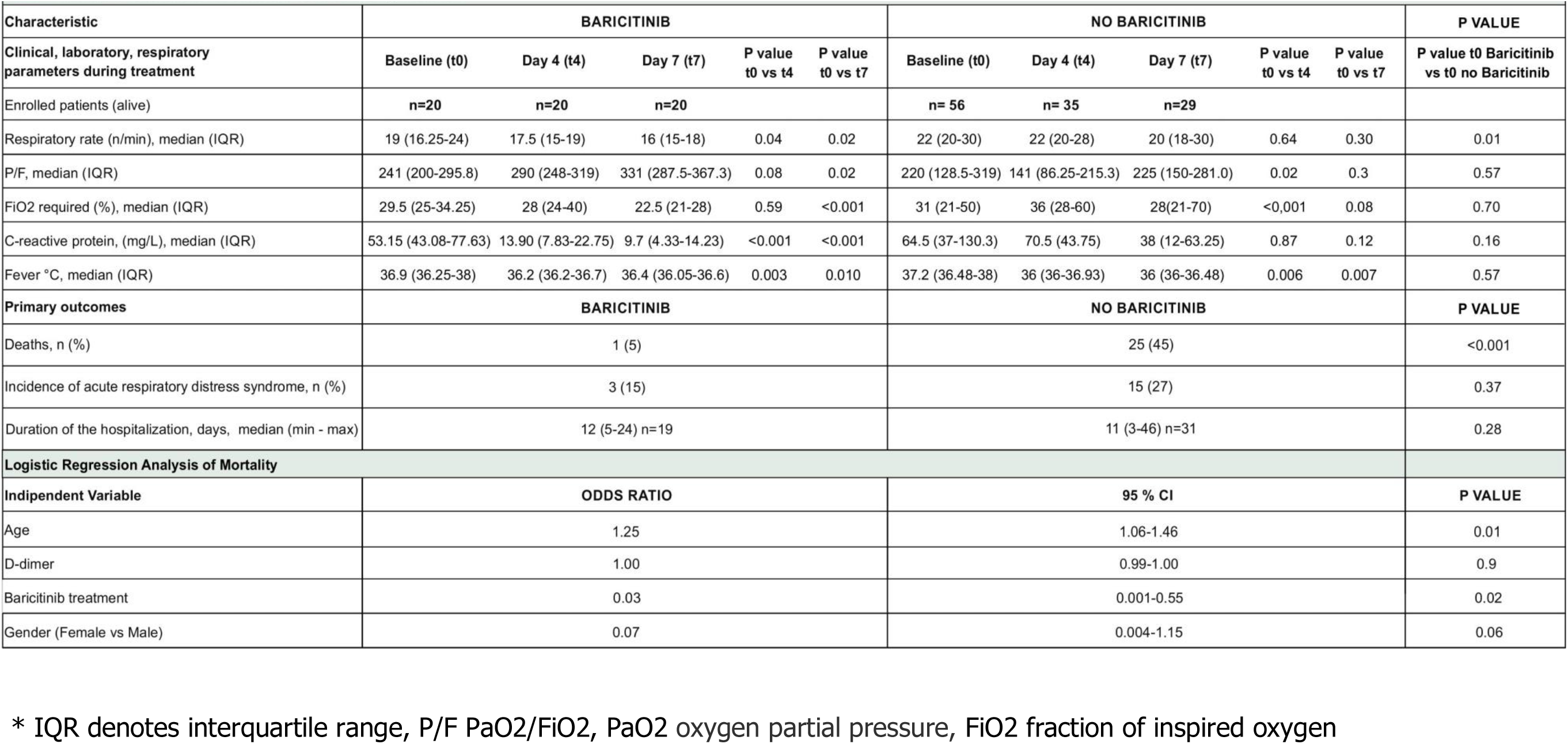
Clinical characteristics of Enrolled Patients During Treatment.

Clinically, the baricitinib-treated patients’ cohort showed a different outcome in terms of mortality. Only one of the 20 baricitinib-treated patients (5%) died after the completion of therapeutic treatment compared to 25 dead patients (45%) out of 56 in non baricitinib-treated patients’ group (p<0.001). In a multiple logistic-regression model adjusted for age, sex and basal D-dimer at the baseline, an association between baricitinib treatment and low mortality was evident. Indeed, age Odds Ratio (OR) was 1.25 (p=0.01), D-dimer OR was 1.00 (p=0.9), baricitinib treatment OR was 0.03 (p=0.02) and gender (female vs. male) OR was 0.07 (p=0.06) (Table 1). On the contrary, we did not observe any significant difference in ARDS incidence or disease duration expressed as time (days) of hospitalization. Finally, to investigate in depth baricitinib impact on resolving COVID-19 pathology, we analyzed the clinical features of baricitinib-treated patients and control at the enrollment (t0) and after four (t4) and seven (t7) days (Table 1). Interestingly, patients treated with baricitinib experienced a faster reduction in oxygen flow need (p<0.001) and a more rapid increase in the P/F ratio compared to the control group (p=0.02), together with a reduction in serum levels of C-reactive protein (p<0.001), whereas no differences in fever resolution were observed between the two groups. (Table 1). When we considered the interstitial lung involvement, either chest X-ray or high resolution computed tomography (HRCT) documented variable extension at different disease stages. We observed an increased interstitial involvement in the control group at the 4^th^ day and a reduction of the same at the 7^th^ day in both groups without statistically relevant differences. Since at the admission time the interstitial lung involvement was more frequent and extensive in patients enrolled in baricitinib group compared to patients in the control cohort, the possible clinical benefit on the lung induced by baricitinib treatment was speculated (data not shown). Collectively, these data demonstrated the impact of a short-term treatment with baricitinib in regularizing the immune landscape in COVID-19 patients. Since this treatment can be orally given to the patients outside the hospitals, the impact in limiting the negative consequences of SARS-CoV-2 during the pandemic spread might be of outmost relevance for the global health care system. The reduced mortality and the clinical benefit in patients treated with baricitinib will have to be established in the ongoing, randomized clinical trials. However, these effects are seemingly not influenced by the presence of higher D-dimer values (a negative prognostic factor) (10) in the non-baricitinib group at baseline, as suggested by the logistic regression analysis.

### Baricitinib affects the immune landscape in COVID-19 patients

In order to evaluate the downstream molecular targets of baricitinib activity, we first demonstrated that patients with COVID-19-related pneumonia expressed phosphorylated Tyr705 in STAT3 (p-STAT3) in different leukocyte subsets (Supplemental Figure 1A). Instead, we were unable to detect p-STAT1 (Tyr701) in these cells (Supplemental Figure 1A). While the viral load should have triggered type I IFN response, which relies on STAT1 signaling, the use of an antibody restricted to p-Tyr701 and not detecting p-Ser727 could have limited the ability to follow STAT1 changes in the samples. Therefore, p-STAT3 was selected to monitor the on-target effect of the drug *in vivo*. Indeed, baricitinib administration produced a significant inhibition of p-STAT3 in T lymphocytes (Supplemental Figure 1B), NK cells (Supplemental Figure 1C), monocytes (Supplemental Figure 1D) and neutrophils (Supplemental Figure 1E), as evaluated in 6 patients. We detected a significant contraction in STAT3 phosphorylation already 4 days following the drug administration, suggesting that treatment achieved an effective on-target activity. Conversely, no statistically relevant activity was observed in B cells during the treatment (Supplemental Figure 1F).

We then analyzed different immune cell populations in the blood of patients who received or not baricitinib. In 12 baricitinib-treated patients we did not detect any modification in the absolute number of circulating leukocytes (Supplemental Figure 2A) as compared to control group (n=8). These patients presented the same clinical features at baseline except for median P/F (p=0.04) and LDH (p<0.01), which were respectively lower and higher in baricitinib treated patients (data not shown). Remarkably, all baricitinib-treated patients showed an increment in absolute number of circulating lymphocytes during the time frame of the treatment, reaching the normal range (1200-2000 cell/µl) by the end of the treatment (t7) (Figure 1A). Interestingly, baricitinib increased the number of circulating T (Figure 1B) and B cells (Figure 1C); in particular, we could observe a significant effect of the drug on circulating CD4^+^ T cells (Figure 1D) and, among them, lymphocytes with an effector-memory phenotype (CD3^+^CD4^+^CD45RA^-^CD27^-^) were particularly expanded (Figure 1E). On the other hand, the absolute number of CD8^+^ T lymphocytes was similar and below the lower range both in baricitinib-treated and control groups, with a tendency to reach the physiological range only in baricitinib-treated patients (Figure 1F). Baricitinib did not affect the absolute number of NK cells (Supplemental Figure 2B) and neutrophils (Supplemental Figure 2C). Notably, in both untreated and baricitinib-treated patients, there was an expansion of monocytes at day 7, which probably reflects a common COVID-19 evolution (Supplemental Figure 2D), as recently published (11). By t-distributed stochastic neighbor embedding (t-SNE) analysis, we confirmed the increment in naïve (from 11.2 % to 13.8 %) and central memory (from 11.9 % to 16.7 %) CD4^+^ T populations and B lymphocytes (from 11.8 % to 15.7%) but also unveiled a shift among the CD8^+^ T cells after baricitinib treatment. Specifically, there was a time-dependent decrease in senescent (CD8^+^CD45RA^+^CD57^+^CD27^-^ cells; from 7.3 % to 3.3 %) with a concomitant increase in both naïve (CD8^+^CD45RA^+^CD57^-^ CD27^+^, from 4.3 % to 5.3 %) and memory (CD3^+^CD8^+^CD27^+^CD45RA^-^, from 3.4 % to 4.8 %) CD8^+^ T lymphocytes, suggesting an effect of baricitinib in supporting effector T cell activation (Figure 1G). To validate our t-SNE analysis, each marker was extracted using functions from the flowCore (Supplemental Figure 3A and B). Conversely, we did not detect changes in the number of HLA-DR^+^CD38^+^ (activated), CD3^+^CD8^+^ T cells (12). It remains to be determined whether the CD8^+^ T cell function and/or repertoire might be altered by the treatment.

**Figure 1:**
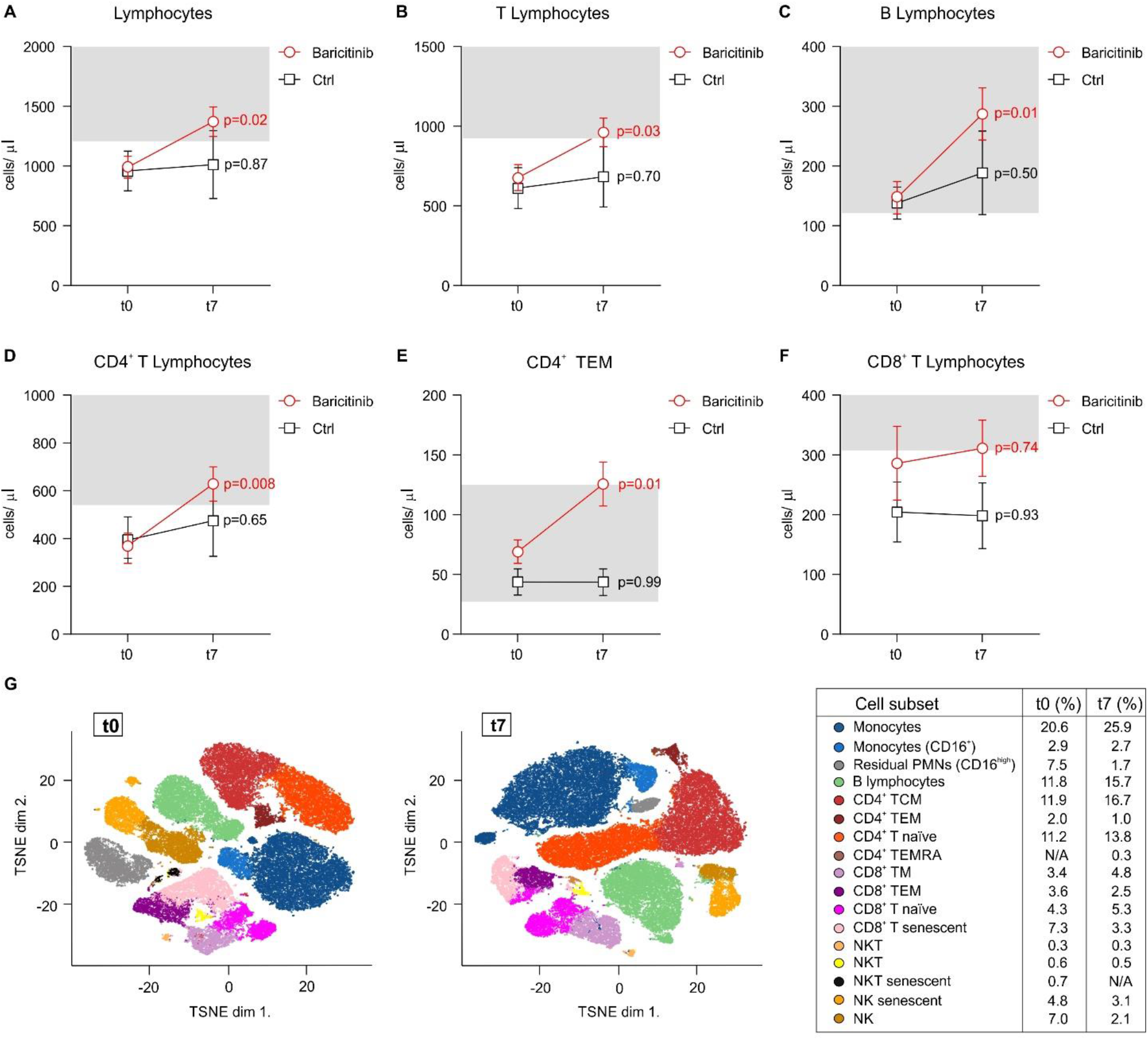
Baricitinib treatment restores normal lymphocyte counts in the blood. Peripheral blood of COVID-19 patients enrolled in either baricitinib (n=12) or basic treatment (n=8, Ctrl) arms was analyzed at t0 (baseline), and t7 (7 days following treatment) by flow cytometry. Number of cells/µl was reported for lymphocytes (A), T lymphocytes (B), B lymphocytes (C), CD4^+^ T lymphocytes (D), CD4^+^ T effector-memory (E), CD8^+^ T lymphocytes (F). Normal reference range is shown in light gray boxes. Data are reported as mean ± SEM. Statistic performed by One-way RM ANOVA. (G) t-SNE analysis of peripheral blood from 12 patients at t0 (left) and t7 (right) of baricitinib treatment. The identified clusters are reported in different colors as follow: monocytes, monocytes (CD16^+^), residual PMNs (CD16^high^), B lymphocytes (CD19^+^CD45RA^+^), CD4^+^ T central memory (TCM, CD3^+^CD4^+^CD27^+^CD45RA^-^), CD4^+^ T effector memory (TEM, CD3^+^CD4^+^CD57^+^CD27^-^CD45RA^-^), CD4^+^ T naïve (CD3^+^CD4^+^CD27^+^CD45RA^+^), CD4^+^ T effector memory re‐ expressing CD45RA (TEMRA, CD3^+^CD4^+^CD45RA^+^CD57^+^), CD8^+^ T memory (TM, CD3^+^CD8^+^CD27^+^CD45RA^-^), CD8^+^ T effector memory (TEM, CD3^+^CD8^+^CD45RA^-^ CD57^+^), CD8^+^ T naïve (CD3^+^CD8^+^CD27^+^CD45RA^+^), CD8^+^ T senescent (CD3^+^CD8^+^CD57^+^CD45RA^+^), NKT (CD3^+^CD16^+^CD56^+^CD45RA^+^), senescent NKT (CD3^+^CD16^+^CD56^+^CD45RA^+^CD57^+^), NK (CD16^+^ CD56^+^CD45RA^+^) and senescent NK (CD16^+^CD56^+^CD45RA^+^ CD57^+^).

Considering the variation in B lymphocyte numbers (Figure 1C), we also evaluated the plasma levels of IgA and IgG specific for the receptor-binding domain (RBD) domain of the SARS-CoV-2 spike protein. As shown in figure 2A and B, while we did not observe a baricitinib-specific effect on the IgA levels between t0 and t7, a significant increase in the IgG was present only in the baricitinib-treated group. Among those who did not present any virus-specific IgG at t0 in this group (n=20), 8 out of 9 developed high titers at t7. The single patient who never presented virus-specific IgG was the only death in the baricitinib group. Instead, in the control group (n=8), among the 7 patients who did not present any virus-associated IgG at t0, only 3 developed virus-specific IgG. Additionally, at t7 the mean level of IgG in the baricitinib-treated group was almost twice than control group.

**Figure 2:**
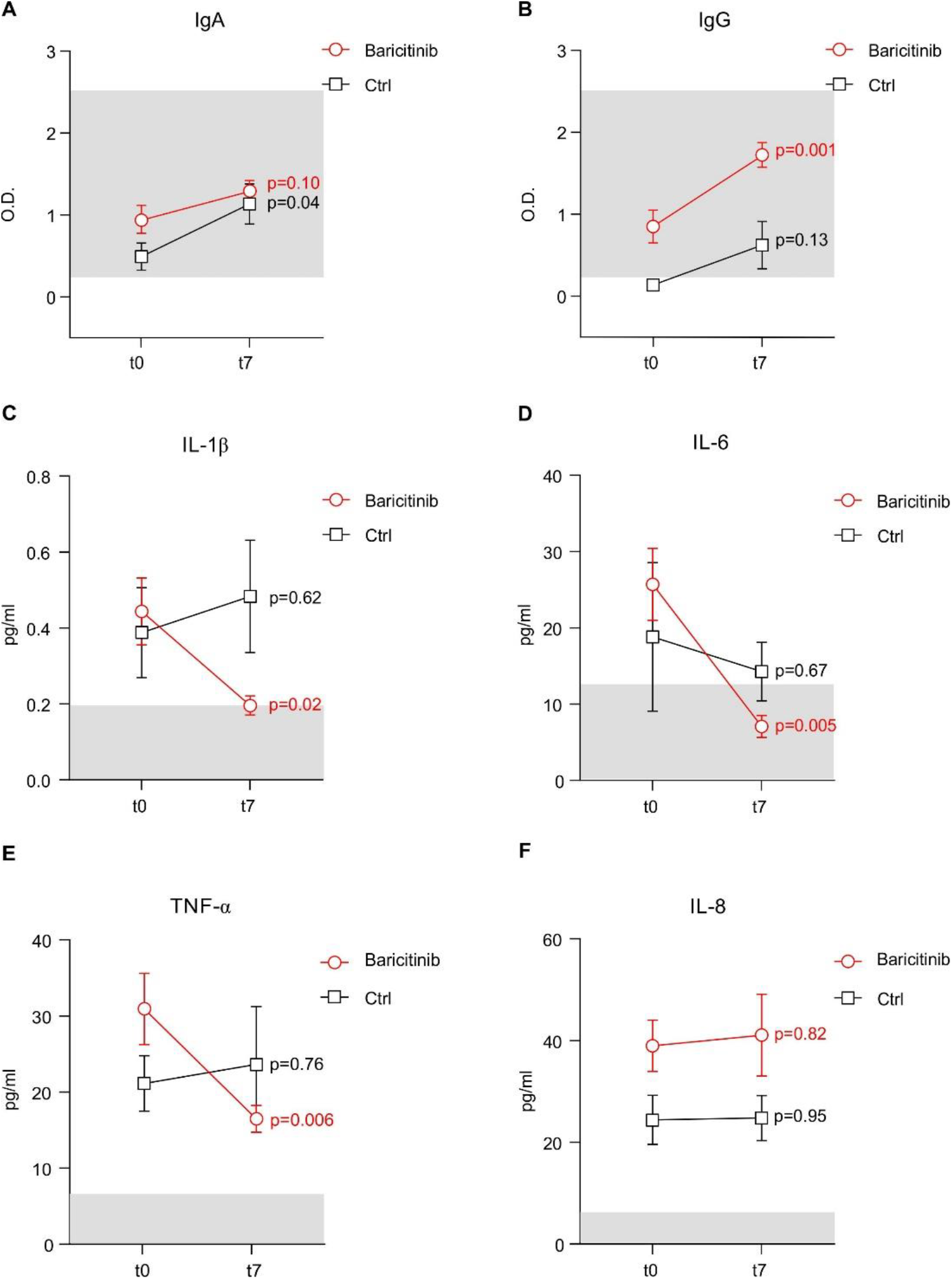
Baricitinib treatment affects IgG levels and production of inflammatory cytokines that contribute to the cytokine storm. Plasma of COVID-19 patients enrolled in either baricitinib (n=20) or basic treatment (n=8, Ctrl) arms was analyzed at t0 (baseline), and t7 (7 days following treatment) to evaluate the concentration of IgA (A), IgG (B), IL-1β (C), IL-6 (D), TNFα (E), and IL-8 (F). For serological data, the light gray boxes identify the range of Ab detection. Normal median value of cytokines is shown by light gray boxes. Data are reported as mean ± SEM. Statistic performed by One-way RM ANOVA.

We then demonstrated that baricitinib treatment normalized the plasma concentration of several pro-inflammatory cytokines, which are produced in abnormal levels in COVID-19 as well as in CRS patients; indeed, 7 days after the first dose we detected a significant reduction in IL-1β, IL-6 and TNFα plasma concentration within the baricitinib-treated patients but not in control group (Figure 2C-E), supporting the standpoint of baricitinib as an effective therapeutic tool against the cytokine storm, a major cause of ARDS and multiple organ failure in COVID-19 patients (4). Interestingly, we did not appreciate any difference in the concentration of IL-8, indicating that JAK-1/JAK-2-dependent molecular pathways are not the main regulators of IL-8 production, at least in these patients (Figure 2F).

### Baricitinib modifies immune suppressive features of myeloid cells

While the efficacy of baricitinib treatment was assessed as a decrease in the intensity of p-STAT3 and levels of pro-inflammatory cytokines, we hypothesized that these alterations might also impact the ability of myeloid cells to modulate T cell proliferation. To verify this aspect, we studied in greater detail the case of a 68 years old woman, admitted to the hospital for the persistence of fever and dyspnea. Lung X-ray analysis revealed a bilateral and interstitial pneumonia, compatible with a positivity to SARS-CoV-2, further confirmed by an oropharyngeal swab. The patient presented a rapidly worsening clinical course, which caused her admittance to the intensive care unit (ICU). Upon hospitalization, she agreed to receive antiviral standard care treatment and off-label baricitinib, which were also continued in ICU. After one week of permanence in ICU, the patient begun to breath spontaneously and was transferred to the pneumology unit. Ten days later, she finally left the hospital in good clinical conditions. From the peripheral blood of the patient we either isolated monocytes (CD14^+^ cells), low and normal density neutrophils (LDNs and NDNs, respectively) at two different moments of hospitalization: during ICU stay (ICU) and when she exited this unit (No ICU). At the same time, we evaluated the serum levels of several pro-inflammatory cytokines and observed a decrease in IL-1β and TNFα levels from the beginning of the treatment (t0) towards the end (t7). Interestingly, IL-6 levels increased at t4 but dropped completely at t7, while IL-8 levels had a tendency to increase during the same time window (Figure 3A). We then evaluated the capacity of the isolated myeloid cells to suppress the proliferation of activated T cells. As shown in figure 4B, the suppressive activity of monocytes (CD14^+^ cells), as well as of their supernatants, decreased when the patient exited the ICU, while it was maintained in CD66b^+^ LDNs on a per cell basis; as expected, the CD66b^+^ NDN fraction was poorly suppressive. While the total count of monocytes seemed to be unaffected by the treatment, we observed an opposite trend in the distribution among monocyte subsets, defined as classical (CD14^high^ CD16^low/dim^) and non-classical (CD14^low/dim^ CD16^high^), with the formers decreasing and the latest increasing during the analyzed time points (Figure 3C). Similarly, baricitinib treatment did not alter the total count of neutrophils, but shifted the distribution from the LDN fraction, which dropped during the treatment, and NDNs, which instead increased (Figure 3D). Of note, when we assessed the cytokine content in the conditioned media obtained from the immune suppressive populations, i.e. monocytes and LDNs analyzed in figure 3B, we noticed that overall monocytes secreted more cytokines than LDNs and that, on a per cell basis, the breadth of cytokine release was generally higher in monocytes (Figure 3E), consistent with published data about the monocyte contribution to the cytokine storm (13, 14).

**Figure 3:**
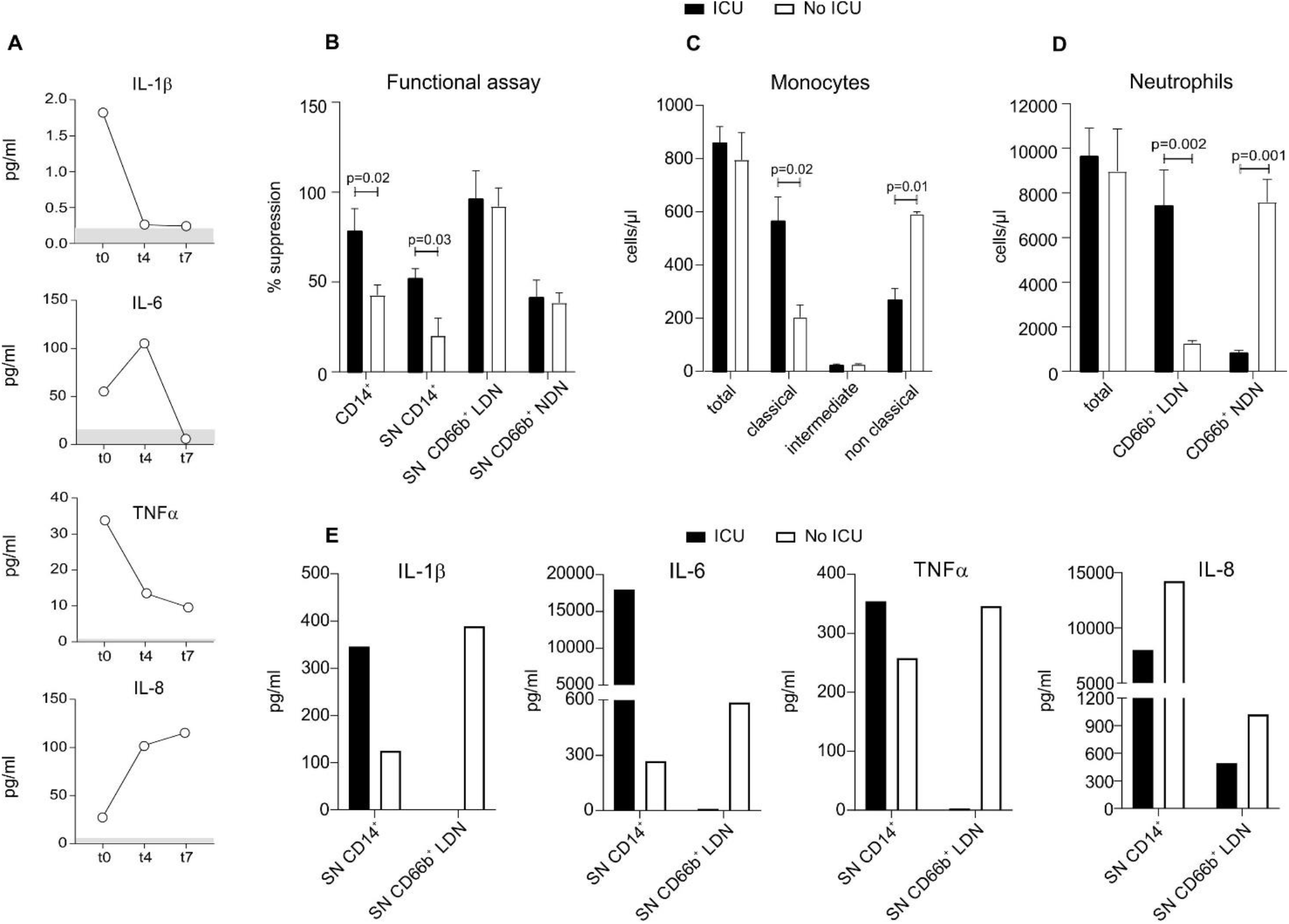
Baricitinib treatment alters the immune suppressive abilities and the distribution of myeloid cells during the recovery phase. The plasma of a COVID-19 patient enrolled in baricitinib arm, who entered ICU during treatment, was analyzed at t0 (baseline), t4 (4 days following treatment) and t7 (7 days following treatment) to assess proinflammatory cytokines (A) by automated immunoassay system. Peripheral blood of the same patient was tested while in ICU and after leaving ICU (No ICU). (B) Monocytes (CD14^+^), low density (CD66b^+^ LDN) and normal density (CD66b^+^ NDN) neutrophils were isolated from the peripheral blood; the immune suppressive abilities of either enriched cells or their conditioned media were tested in functional assay on activated T cells and shown as percentage (%) of suppression. Monocyte (C) and neutrophil (D) subsets were evaluated by flow cytometry and reported as number of cells/µl. (E) Cytokines released in the conditioned media by CD14^+^ and CD66b^+^ LDNs were quantified by multiplex ELISA. (B-D) Data are reported as mean ± SEM.

Collectively, our findings advocate that there might be a substantial advantage in targeting STAT3 pathway. Many viruses might have developed strategies to trigger STAT3 signaling to dampen antiviral innate immune response during the acute phase response, either preventing interferon responses or triggering the negative immune regulatory effects of IL-6 and IL-10 (12, 15). Indeed, STAT3 pathway is relevant for the production of some cytokines during the CRS, including but not limited to IL-6 and IL-10 (16). It is important to highlight how baricitinib also affects the level of IL-1β and TNFα, which are traditionally considered under transcriptional regulation by molecular pathways distinct from JAK1/JAK2, i.e. inflammasome and NFAT/NF-κB, respectively (17-19). From the immune standpoint, a reduction in STAT3 in NK cells promotes a consistent increase in perforin and granzyme B, improving the NK-mediated surveillance against pathogens (20). On the other hand, preventing STAT3 phosphorylation in monocytes and neutrophils affects the ability to produce and release pro-inflammatory cytokines (16) as well as their immunosuppressive properties on T lymphocytes (21). Moreover, STAT3 in cytotoxic CD8^+^ T cells controls lymphocyte differentiation from effector to long-term central memory phenotype (22).

In the context of the health crisis in Northern Italy, where the hospital capacity was quickly overwhelmed by a load of seriously affected patients, the choice of baricitinib was based on several considerations, also of practical value. The first one did not only rely on pathogenesis considerations, but also on the current availability of the drug for RA therapy. Although off-label, baricitinib administration in humans was already authorized by regulatory Authorities for RA thereby facilitating its immediate use. A second consideration was related to pharmacokinetics and pharmacodynamics of the drug. Since early intervention on the cytokine cascade might prevent the progression to virus-induced damage, we anticipated the drug effect by giving a loading dose. Phase I pivotal studies on healthy volunteers, taking standard daily doses of 4 mg of baricitinib, demonstrated that the achievement of plasma steady-state concentration of the drug normally occurs in 48 hours (23). To anticipate this concentration in the first 24 hours, a double dose (4 mg every 12 hours for 48 hours) was planned, followed by the standard dose on the following days. Upon reaching the steady-state concentration, the inhibition of STAT3 phosphorylation occurs in 2-4 hours (23). In this way, the action of the drug on STAT3 target should already be guaranteed within the first 24 hours. The present data confirm that an early effect was indeed reached in T lymphocytes, NK cells, monocytes and neutrophils (Supplemental Figure 1). A third consideration regarded the safety profile of baricitinib (7, 8). This aspect was of crucial importance in the context of an off-label use of the drug.

## Methods

### Patients

Within the period from March 25^th^ to April 18^th^ 2020, patients admitted to the Unit of Internal Medicine at the University Hospital of Verona and Pederzoli Hospital of Peschiera with diagnosis of COVID-19 pneumonia, confirmed by the positivity of nasal swab for SARS-CoV-2 tested with reverse-transcriptase-polymerase-chain-reaction assay, were consecutively enrolled to the study.

The study was designed as an observational longitudinal trial and approved by the local ethical committee (Prot. n° 17963, P.I. Vincenzo Bronte); informed consent was obtained from all the participants to the study. The ClinicalTrials.gov identifier of this project is protocol NCT04438629. All clinical investigations have been conducted according to Declaration of Helsinki principles. A group of subjects (n=20) was treated with baricitinib according to an off-label use of the drug. These patients were not considered for the randomized, multi-center clinical trial that will start recruiting. The use of the drug has been proposed on the basis of a therapeutic protocol with stringent inclusion criteria. In particular, a clinical onset of symptoms not exceeding 9 days and the presence of interstitial lung involvement not exceeding 50% at chest X-ray or computed tomography (CT) were required for the enrollment for baricitinib therapy. The exclusion criteria included the presence of active malignancies and/or immunodeficiency, cardiovascular disease with recent myocardial infarction or stroke, as well as thrombophilia or deep venous thrombosis or pulmonary thromboembolism. Moreover, the presence of chronic kidney disease with renal failure, cirrhosis Child Pugh C or the presence of anemia or severe neutropenia or lymphocytopenia were listed as exclusion criteria (Supplemental Table 1).

In total, 88 patients (44 M/44 F) affected by COVID-19-related pneumonia were followed during the hospitalization to analyze different clinical outcomes. All subjects were treated with either hydroxicloroquine or antiviral therapy (lopinavir/ritonavir) as single agents or in combination (hydroxicloroquine plus antiviral therapy) according to clinical features. Supportive therapy, such as antibiotic prophylaxis and anticoagulant treatment, was provided at the discretion of the clinicians (Table 1). Steroids therapy was systematically avoided. Twelve (6 M/6 F) of these patients were excluded from the analysis considering their positive active history of malignancies: 2 hematological disorders (a multiple myeloma and an acute myeloid leukemia) and 10 cases of solid malignancies, including lung and breast cancers as well as kidney, prostate, ovarian and gastro-intestinal tumors. Only 3 out of these 12 subjects received chemotherapy at the time of infection while, the remaining subjects, were off therapy according to the disease’s stage. Arterial hypertension and cardiovascular disease as well as diabetes, chronic obstructive pulmonary disease, and chronic kidney disease were the prevalent pathologies in the other 76 subjects (Supplemental Table 1). Among them, 20 subjects received the full course of baricitinib according to the study protocol. The other 56 subjects were considered as control group.

According to the inclusion criteria and to baricitinib pharmacokinetics, patients were treated with baricitinib 4 mg twice daily for 2 days followed by 4 mg per day for the remaining 7 days. A low dose of 2 mg twice daily for 2 days followed by 2 mg per days was maintained for patients older than 75 years of age. A dose reduction was also considered in case of renal insufficiency (GFR < 30 ml/min/1.73 m^2^), hepatotoxicity or myelotoxicity.

Clinical features during treatment were recorded for all patients included in the study. Flow cytometry, cytokine and serology assays were performed in a subgroup of patients on the basis of biological sample availability.

### Study assessment

This off-label treatment was evaluated using laboratory values, including serum concentration of C-reactive protein (PCR) and oxygenation index (PaO2/FiO2, P/F), as well as immunological parameters, including serum cytokine level (IL-6, IL-1β, TNFα and IL-8), serology and quantification of absolute number of different immune cell populations from the enrollment to day 7. Further details regarding flow cytometry, cell purification, ELISA, and cellular assays are provided in the Methods section of the Supplementary Appendix. We quantified the incidence of key clinical elements such as oxygen flow need, P/F ratio, radiology score calculated on the basis of the percentage of interstitial lung involvement (score 0=no interstitial lung involvement; score 1 < 20% of interstitial involvement; score 2= parenchymal involvement included between 20 and 50%; score 3= parenchymal involvement included between 50 and 70%; score 4 in case of interstitial involvement > 75%), respiratory rate as well as ARDS incidence, duration of hospitalization, and death.

### Statistical analysis

The clinical analysis (highlighted in Table 1 and 2) at the baseline (t0) included all patients enrolled in the study for whom laboratory tests were available. Given the nature of this off-label program, there were some missing data. For analysis during the treatment we considered only those patients who were still alive, for whom the clinical and laboratory data for at least two time points were available. Analysis on the duration of hospitalization was performed in all the patients enrolled except for the ones who died. Quantitative variables were expressed as the median and interquartile range (IQR), qualitative ones as percentages. To perform pairwise comparisons, significance of difference was evaluated by the Mann-Witney U-test for quantitative variables and Fisher’s exact test for categorical variables. The Wilcoxon matched-pairs signed-rank test was used to test the equality of matched pairs of observation. Multiple logistic-regression analysis was performed to assess the association between the treatment and mortality for known negative prognostic factor (age, sex and D-dimer) (10). The strength of the association was expressed by the Hazard Ratio (HR) with 95% confidence interval (CI). Statistical significance was set at p-value <0.05, and clinical analyses were performed using statistical software STATA^®^ version 16.0 (StataCorp, College Station, TX, USA). For statistical laboratory analyses we performed paired comparisons by Student’s t-test and one-way Anova test for repeated measures, using Graph Pad Prism (San Diego, California, version 8.4.2).

## Data Availability

The authors declare that all the other data supporting the findings of this study are available within the article and its supplementary information files and from the corresponding authors upon reasonable request.

## Author contributions

V.B., C.L. and O.O. designed the research; V.B. and S.U. coordinated the study; E.T., V.B., E.P., K.D., S.F., F.P., D.R., P.B., M.R., A.C., W.M., C.L. and O.O. collected clinical specimens and data; L.T. performed the statistical analysis; A.V., F.D.S., R.T., A.F., L.P., C.M. and S.C. completed flow cytofluorimetric acquisition and analysis; V.P., F.H. and M.I. tested serum and plasma cytokines; S.C., R.M.B. did functional analysis; F.F. and P.P. performed serological analysis and prepared key reagents; S.U., E.T., F.D.S., R.T., and S.C. analyzed the results; S.U., E.T., S.C., O.O. and V.B. wrote the manuscript.

## Acknowledgement

This work was supported by Fondazione Cariverona (ENACT Project) and Fondazione TIM.

We thank all patients who participated in this study and their families. We also thank all the personnel involved in patients care and assistance. We thank the members of Immunology Section of Verona University Hospital who actively worked during the pandemia: Morena Martini, Fiorenza Paiola, Elena Lucchini, Claudia Pizzoli, Elena Chiesa, Oretta Gabrielli, Nadia Brutti, Monica Brentegani, Elisabetta Gallo, Giulio Fracasso, Tiziana Cestari, Ornella Poffe and Cristina Anselmi for the excellent technical work; Cristina Frusteri, Giovanna Zanoni, Silvia Sartoris, Riccardo Ortolani and Selena Gomirato for their help with the management of immunological data; Daniel Lovato, Antonella Valentini and Claudia Italia for the administrative support.

We dedicate this work to the memory of health care workers who have given their lives in the care of patients with COVID-19.

## Conflict of interest

The authors declare no competing interests.

## Notes

### Competing Interest Statement

The authors have declared no competing interest.

### Clinical Trial

NCT04438629

### Author Declarations

The study was approved by the Institutional Review board of Azienda Ospedaliera Universitaria di Verona Prot. n.17963 (25 March 2020)

